# Provider unit costs and cost drivers of in vitro fertilization (IVF) services in a public tertiary hospital in Tanzania: A micro-costing study

**DOI:** 10.64898/2026.10.20.26365987

**Authors:** Godwill Einhard Sanga, Mohamed Zahir Alimohamed, Alphoncina Kagaigai, Phares G. Mujinja

## Abstract

**Background:** Infertility affects up to 32.0% of couples in sub-Saharan Africa, while access to assisted reproductive technologies (ART), including In Vitro Fertilization (IVF), remains limited by high treatment costs. As Tanzania advances Universal Health Coverage (UHC) through the National Health Insurance Act of 2023, empirical provider cost estimates are needed to inform reimbursement and strategic purchasing. This study estimated the provider unit cost, cost structure, and cost drivers of IVF services at Muhimbili National Hospital (MNH), Tanzania.

**Methods:** A provider-perspective cross-sectional micro-costing study was conducted at MNH between January and December 2025. Bottom-up micro-costing was applied to direct costs, while top-down allocation was used for capital and indirect costs. All 10 IVF cycles initiated during the study period were included, comprising five completed and five incomplete cycles. Deterministic one-way sensitivity analysis assessed the robustness of cost estimates.

**Results:** The total provider cost for five completed IVF cycles was TZS 55,410,375.00 (USD 22,164.15), corresponding to a mean unit cost of TZS 11,082,075.00 (USD 4,432.83) per cycle. Direct costs accounted for 99.92% of the total cost, while indirect costs contributed 0.08%. Monitoring, investigation and ovarian stimulation was the largest treatment-stage cost component (55.13%). Among direct variable costs, drugs and hormones accounted for 52.67%, followed by reagents and media at 33.05%; together, these accounted for 85.72% of direct variable costs and represented the principal recurrent cost drivers. Sensitivity analysis showed that varying drugs and hormones by ±15% changed the estimated provider unit cost from TZS 10,474,000 (USD 4,189.60) to TZS 11,690,125 (USD 4,676.05), while variation in reagents and media changed it from TZS 10,700,550 (USD 4,280.22) to TZS 11,463,600 (USD 4,585.44).

**Conclusion:** IVF service delivery at MNH was predominantly driven by direct clinical resource inputs. Drugs, hormones, reagents and media were the principal recurrent cost drivers. These findings provide empirical evidence to support NHIF reimbursement, strategic purchasing, and resource allocation for publicly financed IVF services in Tanzania.

## Introduction

Infertility is a disease of the reproductive system characterized by the failure to achieve a clinical pregnancy after 12 months or more of regular unprotected sexual intercourse [1]. Globally, an estimated 17.5% of adults experience infertility, with the burden reaching up to 32.0% in sub-Saharan Africa [2]. In Tanzania, infertility remains an important public health concern, driven by reproductive tract infections, delayed diagnosis, and limited access to specialized fertility services despite a total fertility rate of 4.8 births per woman [3].

In Vitro Fertilization (IVF) is the principal assisted reproductive technology for managing tubal, male factor, and unexplained infertility [4]. However, IVF services in sub-Saharan Africa have largely been confined to high-cost private facilities, limiting access for many households [2]. Tanzania has expanded public provision of IVF services through Muhimbili National Hospital (MNH), yet provider reimbursement and pricing have largely been established without empirical provider cost estimates [4,5].

The implementation of Tanzania’s National Health Insurance Act of 2023 has renewed the need for evidence on the cost of specialized health services to support Universal Health Coverage (UHC) [5]. Provider unit cost estimates are essential for informing reimbursement, tariff setting, and strategic purchasing, particularly for resource-intensive interventions such as IVF [6]. Despite this policy need, evidence on the provider cost of public IVF services in Tanzania remains limited.

This study estimated the provider unit cost, cost structure, stage-specific cost distribution, cost drivers, and cost uncertainty of IVF services at Muhimbili National Hospital using a provider-perspective micro-costing approach. The findings provide empirical evidence to inform reimbursement decisions and strategic purchasing of IVF services within Tanzania’s evolving UHC framework [5,6].

## Methods

### Study Site

The study was conducted at the In Vitro Fertilization (IVF) and Assisted Reproductive Technology (ART) Centre of Muhimbili National Hospital (MNH) in Dar es Salaam, Tanzania [3]. Muhimbili National Hospital is the national tertiary referral and teaching hospital with a bed capacity of 1,500 and serves as the principal teaching hospital for the Muhimbili University of Health and Allied Sciences (MUHAS) [3,9]. The hospital was purposively selected because it hosts Tanzania’s first public-sector IVF and ART centre, equipped with specialized embryology laboratories and multidisciplinary expertise for fertility care [3,9,13]. As the country’s leading public provider of assisted reproductive services, the centre offers an appropriate setting for estimating provider unit costs and resource utilization under routine clinical practice. IVF services are financed through a combination of public and private health insurance schemes and out-of-pocket payments by clients [5].

### Study Design

A hospital-based cross-sectional descriptive economic evaluation was conducted from the healthcare provider perspective to estimate the unit cost of delivering In Vitro Fertilization (IVF) services at Muhimbili National Hospital [4,6]. The analysis captured resource utilization and costs associated with routine IVF service delivery over a one-year study and analytical horizon, from 1 January to 31 December 2025 [8,14].

### Data Collection Method

A bottom-up micro-costing approach was used to collect quantitative resource utilization and cost data from patient electronic medical records, embryology laboratory logs, and hospital financial records [4,10]. Resource quantities were obtained from patient records and laboratory registers, while unit prices for pharmaceuticals, culture media, laboratory reagents, and medical consumables were obtained from the Medical Stores Department (MSD) price catalogue and institutional financial records [5,12]. Data extracted included IVF treatment stage, medications, laboratory investigations, clinical procedures, medical consumables, personnel involved in service delivery, and treatment duration for each patient cycle [4,13]. All costs were recorded in Tanzanian Shillings (TZS) and converted to United States Dollars (USD) using the official exchange rate for the study period (USD 1 = TZS 2,500) [8].

### Study Population

The study population comprised all In Vitro Fertilization (IVF) cycles initiated at the Muhimbili National Hospital IVF and Assisted Reproductive Technology Centre between 1 January and 31 December 2025 (N = 10) [1,2]. A census approach was employed, including all eligible cycles initiated during the study period. Of the 10 initiated cycles, five were completed through oocyte retrieval, fertilization, embryo culture, and embryo transfer, and were used for provider unit cost estimation. The remaining five incomplete cycles, which were discontinued at different treatment stages due to poor ovarian response or fertilization failure, were included to describe stage-specific cost accumulation and resource utilization [1,4].

### Sample Size Determination

No formal sample size calculation was undertaken because the study employed a census approach. All 10 IVF cycles initiated at the Muhimbili National Hospital In Vitro Fertilization and Assisted Reproductive Technology Centre between 1 January and 31 December 2025 were identified and assessed. Of these, five completed cycles met the criteria for the primary provider unit-cost analysis, while five incomplete cycles were retained for supplementary stage-specific cost analysis.[9].

### Inclusion Criteria

The study included all IVF cycles initiated at the Muhimbili National Hospital In Vitro Fertilization and Assisted Reproductive Technology Centre between 1 January and 31 December 2025 with complete clinical and financial records in the electronic medical record system. Eligible records contained the information required for provider cost estimation, including IVF treatment stage, medications, laboratory investigations, clinical procedures, personnel involved in service delivery, and resource utilization [1,9].

### Exclusion Criteria

The following records were excluded from the analysis:

- IVF cycles with incomplete clinical or financial information required for cost estimation [9].
- IVF cycles with major coexisting medical conditions that could substantially alter routine resource utilization and provider costs [3,9].

### Data Collection Procedure

Quantitative cost and resource utilization data were extracted from electronic medical records, annual operational reports, institutional financial records, and the Medical Stores Department (MSD) price catalogue using a standardized data extraction tool [5,10,12]. Information on resource quantities, clinical procedures, personnel time, pharmaceuticals, laboratory investigations, medical consumables, and capital resources was collected for each eligible IVF cycle. The extracted data were reviewed for completeness, consistency, and internal agreement across clinical, laboratory, and financial records before being compiled into Microsoft Excel spreadsheets for subsequent cost analysis [5,10]. Records with missing information required for provider cost estimation were excluded according to the predefined eligibility criteria. No imputation was undertaken for missing cost or resource-use data.

### Costing Procedure

Costs were estimated from the healthcare provider perspective using a bottom-up micro-costing approach to identify, measure, and value all resources consumed in delivering IVF services [4,6,10]. Direct cost components included pharmaceuticals, laboratory investigations, clinical procedures, medical consumables, personnel, and capital resources. Resource quantities were measured in natural units and multiplied by their respective unit prices to estimate total costs using the standard micro-costing approach (Total Cost = Quantity × Unit Price) [6,10]. All costs were valued in Tanzanian Shillings (TZS) and converted to United States Dollars (USD) using the official exchange rate for the study period [8].

Personnel costs for medical specialists, embryologists, registrars, and nurses were estimated by converting monthly basic salaries into a cost per working minute and multiplying this by the time spent delivering services for each IVF cycle [6,15]. Unit prices for pharmaceuticals, laboratory reagents, culture media, and medical consumables were obtained from the Medical Stores Department (MSD) price catalogue and verified against institutional procurement records and Tanzania Medicines and Medical Devices Authority (TMDA) reference sources where applicable [5,12].

The unit of analysis was the IVF treatment cycle. Cost centres were defined, resource inputs were identified, measured, and valued, and total input costs were allocated to the corresponding service outputs to estimate the provider unit cost per completed IVF cycle [6,10,14]. Deterministic one-way sensitivity analysis was undertaken to assess the influence of key cost parameters on the estimated provider unit cost [6,7].

Capital equipment, including CO_2_ incubators, micromanipulators, and ultrasound systems, was annualized using the WHO-CHOICE approach with a 3% discount rate and a useful life of seven years [7,11]. Annual equivalent costs were estimated using the following formula: E={K.r}/{1-(1+r)^{-n}} where *E* is the annual equivalent cost, *K* is the replacement cost of the asset, *r* is the discount rate (0.03), and *n* is the useful life of the asset in years [6,7,11.

### Data Analysis

Quantitative data were analysed using Microsoft Excel 365 and Stata [6]. Resource quantities and unit prices were used to calculate costs for individual inputs, which were subsequently aggregated into direct, indirect, and capital costs [6,10]. The total provider cost was divided by the number of completed IVF cycles to estimate the average provider unit cost per cycle [6,14]. Labor costs were valued using applicable public-sector salary scales and scheme-of-service reference documents [5]. Cost components were summarized to identify the principal cost drivers of IVF service costs.

### Ethical Issues

Ethical approval was obtained from the Muhimbili University of Health and Allied Sciences (MUHAS) Institutional Review Board, with permission from the relevant authorities at Muhimbili National Hospital. Authorization was obtained to access facility-level financial, administrative, and service records. Confidentiality was maintained through anonymization and secure, restricted-access storage of study data [9].

## Results

### IVF Cycles Included in the Costing Analysis

Between January and December 2025, 10 IVF cycles were initiated at the Muhimbili National Hospital (MNH) IVF and Assisted Reproductive Technology Centre. Five cycles (50.0%) were completed through the full treatment pathway and constituted the primary denominator for estimating the provider unit cost. The remaining five cycles (50.0%) were incomplete and reached different treatment stages before discontinuation or deferral. The primary cost analysis was therefore restricted to the five completed cycles to ensure a consistent full-cycle costing denominator, while costs incurred by incomplete cycles were assessed separately using a stage-based approach and were not included in the primary unit-cost estimate.

Estimated cumulative provider costs for the incomplete cycles varied according to the highest treatment stage reached, from TZS 6,110,000 (USD 2,444.00) for monitoring and investigation only to TZS 10,890,000 (USD 4,356.00) for embryo culture and transfer preparation. These costs were excluded from the primary unit-cost denominator to maintain a consistent definition of a completed IVF treatment episode ( Fig 1).

**Figure 1.**
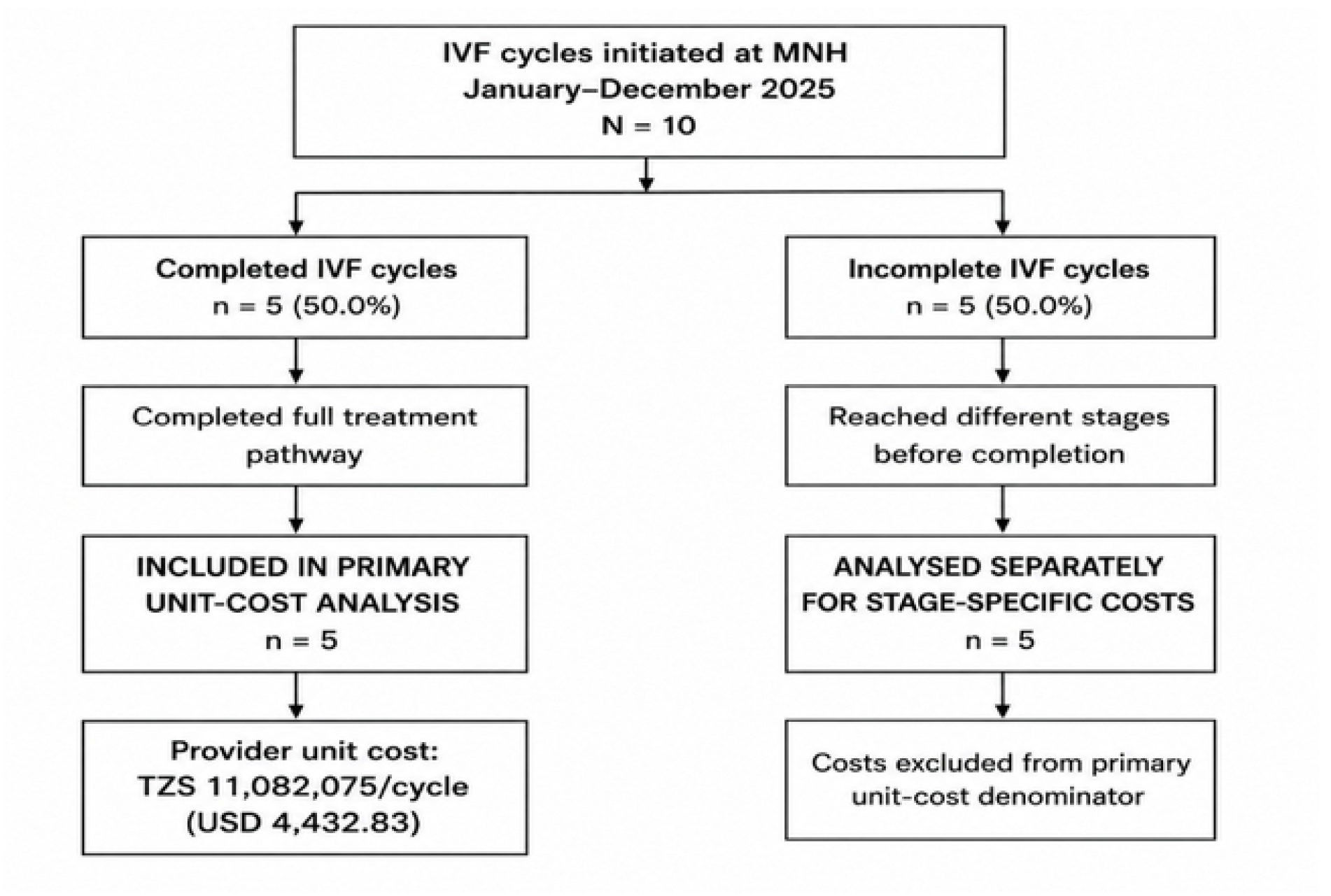
Flow of IVF cycles included in the costing analysis.

**Table 1.** here: Stage-specific cumulative costs of incomplete IVF cycles.

| Incomplete cycle | Highest stage reached | Status | Cumulative cost (TZS) | Cumulative cost (USD) |
| --- | --- | --- | --- | --- |
| IC-01 | Stage 1: Monitoring & investigation | Discontinued | 6,110,000 | 2,444.00 |
| IC-02 | Stage 2: Ovarian stimulation / OPU preparation | Discontinued | 6,760,000 | 2,704.00 |
| IC-03 | Stage 3: Oocyte retrieval & embryology | Deferred | 7,790,000 | 3,116.00 |
| IC-04 | Stage 4: Embryo culture & transfer preparation | Discontinued | 10,890,000 | 4,356.00 |
| IC-05 | Stage 1: Monitoring & investigation | Discontinued | 6,110,000 | 2,444.00 |

### Provider Unit Cost per Completed IVF Cycle

The total provider cost for the five completed IVF cycles was TZS 55,410,375 (USD 22,164.15), corresponding to a mean provider unit cost of TZS 11,082,075 (USD 4,432.83) per completed IVF cycle. Direct costs accounted for 99.92% (TZS 11,073,175; USD 4,429.27) of the unit cost, while indirect costs contributed 0.08% (TZS 8,900; USD 3.56).

Cost accumulation was highest during Stage 1 (monitoring, investigation and ovarian stimulation), which contributed TZS 6,110,000 (USD 2,444.00) per completed cycle. This was followed by Stage 4 (embryo transfer) at TZS 3,100,000 (USD 1,240.00), Stage 3 (embryology) at TZS 1,030,000 (USD 412.00), Stage 2 (ovum pick-up) at TZS 650,000 (USD 260.00), and Stage 5 (luteal support and pregnancy testing) at TZS 192,075 (USD 76.83). Together, these stage-specific costs summed to the total provider unit cost of TZS 11,082,075 (USD 4,432.83).

**Table 3.** Provider unit cost per completed IVF cycle and stage-specific cost distribution at MNH.

| IVF treatment stage | Cost per cycle (TZS) | Cost per cycle (USD) |
| --- | --- | --- |
| Stage 1: Monitoring, investigation and ovarian stimulation | 6,110,000 | 2,444.00 |
| Stage 2: Ovum pick-up (OPU) | 650,000 | 260.00 |
| Stage 3: Embryology | 1,030,000 | 412.00 |
| Stage 4: Embryo transfer (ET/FET) | 3,100,000 | 1,240.00 |
| Stage 5: Luteal support and pregnancy testing | 192,075 | 76.83 |
| **Total provider unit cost per completed cycle** | **11,082,075** | **4,432.83** |

*Note: Stage-specific costs were derived using the stage-accumulation framework and sum to the provider unit cost per completed IVF cycle.*

### Cost Drivers and Sensitivity Analysis

The cost structure was dominated by drugs and hormones, which accounted for 52.67% (TZS 4,053,750; USD 1,621.50) of direct variable costs, followed by reagents and media at 33.05% (TZS 2,543,475; USD 1,017.39). Together, these two categories accounted for 85.72% of direct variable costs, identifying pharmaceuticals and specialized laboratory inputs as the principal recurrent cost drivers of IVF provision.

A one-way deterministic sensitivity analysis was conducted by independently varying major cost components by ±15%, while holding all other parameters constant. Using a base-case provider unit cost of USD 4,432.83 per completed IVF cycle, drugs and hormones produced the greatest change in the unit cost, from USD 4,189.60 to USD 4,676.05. Reagents and media produced the second-largest change (USD 4,280.22–4,585.44), followed by consumables (USD 4,370.60–4,495.06), direct labor (USD 4,389.96–4,475.70), and laboratory investigations (USD 4,429.08–4,436.58).

Thus, the sensitivity analysis identified drugs and hormones as the most influential cost driver, followed by reagents and media, while laboratory investigations had the least influence on the estimated provider unit cost (Fig 2).

**Figure 2.**
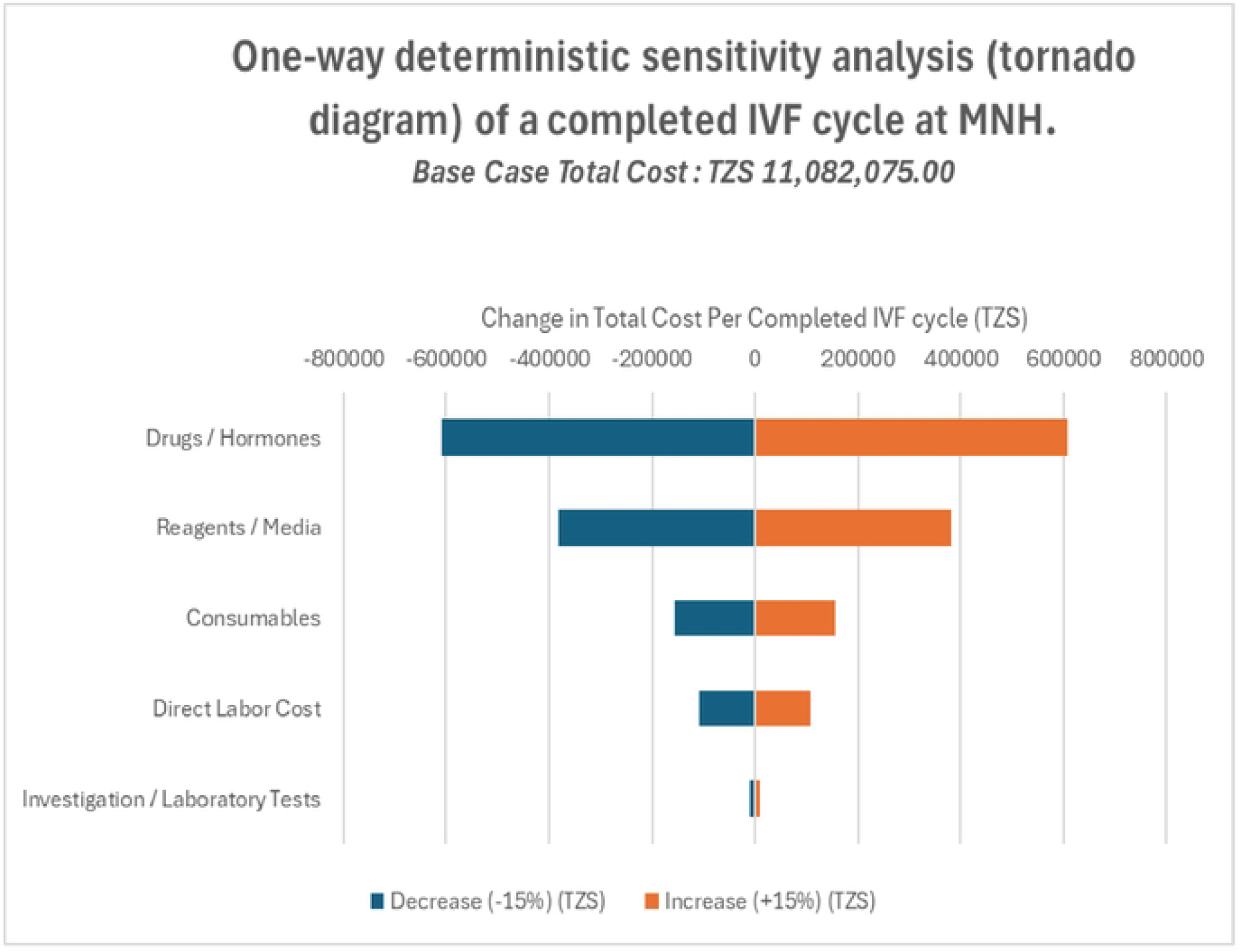
Tornado diagram showing one-way deterministic sensitivity analysis of the provider unit cost.

**Table 4.** One-way deterministic sensitivity analysis of major cost drivers for a completed IVF cycle at MNH.

| Cost component | Lower unit cost (USD) | Base-case (USD) | Upper unit cost (USD) | Absolute impact (USD) |
| --- | --- | --- | --- | --- |
| Drugs/hormones | 4,189.60 | 4,432.83 | 4,676.05 | $\pm 243.23$ |
| Reagents/media | 4,280.22 | 4,432.83 | 4,585.44 | $\pm 152.61$ |
| Consumables | 4,370.60 | 4,432.83 | 4,495.06 | $\pm 62.23$ |
| Direct labor | 4,389.96 | 4,432.83 | 4,475.70 | ±42.87 |
| **Investigation/laboratory tests** | **4,429.08** | **4,432.83** | **4,436.58** | **±3.75** |

*Note: Each parameter was varied independently by ±15%, with all other parameters held constant. The base-case provider unit cost was USD 4,432.83 per completed IVF cycle.*

## Discussion

The provider cost of a completed IVF cycle at Muhimbili National Hospital (MNH) was TZS 11,082,075 (USD 4,432.83), providing an important facility-level benchmark for IVF provision in Tanzania, where detailed provider-cost evidence remains limited. Using a bottom-up micro-costing approach and capital annualization techniques [6, 7, 10], the estimate captures the economic resources required to deliver a completed cycle from the provider perspective and should not be interpreted as a patient charge, reimbursement rate, or national average.

The cost structure demonstrates that IVF provision is predominantly driven by direct clinical inputs. Direct costs accounted for 99.92% of total cost, with drugs and hormones (52.67%) and reagents and media (33.05%) constituting the largest variable components. The substantial contribution of specialized pharmaceuticals and laboratory inputs is consistent with the resource-intensive nature of IVF, where highly specialized clinical and embryology processes require skilled personnel, specialized technologies, and laboratory materials [1, 14–16]. The importance of resource-intensive specialist care is also consistent with previous economic evidence from MNH showing that personnel inputs and active clinical contact time can account for a substantial proportion of provider production costs in specialized tertiary services [3, 10].

The sensitivity analysis further demonstrated the importance of these inputs. Drugs and hormones generated the largest change in unit cost when varied independently by ±15% (±USD 243.23), followed by reagents and media (±USD 152.61) and consumables (±USD 62.23). This indicates that changes in high-value clinical inputs can materially affect the provider cost of IVF and highlights procurement efficiency, accurate forecasting, inventory management, and minimization of wastage as potential areas for cost management. Annualized capital costs also contributed substantially (24.04%; USD 1,064.68), reflecting the capital-intensive nature of IVF [6, 7]. Because annualized capital costs are distributed across completed treatment cycles, the relatively low treatment volume observed during the study may have increased the average cost per cycle. Thus, the observed unit cost should not be interpreted as the long-run marginal cost of an additional IVF cycle.

The incomplete cycles further demonstrate that substantial provider resources are consumed even when treatment does not result in a completed cycle. Although excluded from the primary completed-cycle denominator, their stage-specific costs ranged from TZS 6,110,000 to TZS 10,890,000, with Stage 1 alone accounting for TZS 6,110,000 (USD 2,444). This is economically important because IVF resource use begins well before embryo transfer, with monitoring, ovarian stimulation, laboratory investigations, fertilization/ICSI, and embryo culture requiring substantial clinical and laboratory inputs [1, 14, 15].

Overall, the findings indicate that IVF costs at MNH are primarily determined by specialized clinical inputs, particularly drugs, hormones, laboratory resources, and capital investment, providing useful evidence for provider budgeting, procurement planning, service expansion, and future economic evaluation of IVF financing in Tanzania.

### Limitations

This study has several limitations. First, the primary provider unit-cost estimate was based on only five completed IVF cycles, reflecting the small number of completed cycles available during the study period and limiting the precision and generalizability of the estimate. Second, the study was conducted at a single tertiary public hospital, and the findings may not be directly generalizable to other public or private IVF facilities with different service volumes, input prices, capacity utilization, or organizational arrangements. Third, some shared hospital overheads could not be reliably allocated to IVF using the available financial records, which may have resulted in underestimation of indirect costs. Accordingly, the estimated unit cost should be interpreted as a facility-level estimate under the observed operating conditions rather than a national average.

## Conclusion

This study provides facility-level evidence on the provider cost and cost structure of IVF services at Muhimbili National Hospital. The estimated provider cost of a completed IVF cycle was TZS 11,082,075 (USD 4,432.83), with costs overwhelmingly concentrated in direct clinical inputs (99.92%) [4, 6, 10]. Drugs and hormones and specialized reagents and media were the principal recurrent cost drivers, while annualized capital costs also represented an important component of the cost of service provision [6, 7, 11]. The sensitivity analysis confirmed that changes in drugs and hormones and laboratory inputs have the greatest influence on the estimated unit cost [6, 7, 12]. Overall, the findings demonstrate that IVF is a resource-intensive service in which the observed average cost is influenced by input prices and the relatively low volume of completed cycles. The estimated unit cost therefore provides a useful empirical benchmark for provider budgeting, procurement planning, capacity utilization, and future financing and economic evaluation of publicly provided IVF services in Tanzania [5, 6].

### Recommendations

At the facility level, Muhimbili National Hospital should prioritize efficient management of the major cost-driving inputs, particularly drugs, hormones, reagents, and laboratory media, through improved forecasting, procurement planning, inventory management, and minimization of wastage [5, 10, 12]. The hospital should also strengthen scheduling and workflow coordination to improve utilization of existing IVF infrastructure and specialized personnel [3, 14, 15]. Given the contribution of annualized capital costs [7, 11] and the relatively low number of completed cycles observed, increasing utilization of available capacity, where clinically and operationally feasible, could improve average cost efficiency by distributing annualized costs across a greater number of completed treatment cycles. Monitoring incomplete treatment episodes should also be maintained to identify points of resource loss and opportunities to improve conversion of treatment inputs into completed cycles [4, 13].

At the health-system level, the provider cost estimate should be considered as empirical input for the development and review of appropriate financing, reimbursement, budgeting, and strategic purchasing arrangements for IVF services within Tanzania’s evolving UHC framework [5, 6]. However, the estimate should not be applied as a national tariff without consideration of differences in service volume, input prices, capacity utilization, and institutional arrangements.

Future research should extend this evidence through multi-facility costing studies with larger samples, integrate provider costs with clinical outcomes to support cost-effectiveness analysis [6, 7], and assess household costs and financial consequences of IVF to complement the provider perspective [1, 13]. Such evidence would provide a stronger basis for determining the efficiency, affordability, and broader economic implications of expanding access to IVF within Tanzania’s health system.

## Data Availability

"All relevant data underlying the findings of this study are available within the manuscript and its supporting information files. Due to institutional data governance policies of Muhimbili National Hospital (MNH) and restrictions regarding proprietary hospital financial and procurement records, the raw operational and pricing datasets are available from the corresponding author (Godwill Einhard Sanga) upon reasonable request for researchers who meet the criteria for access to confidential data."

## Supporting information

- **S1 Fig**. Flow of IVF cycles included in the costing analysis.
- **S2 Fig**. Tornado diagram showing one-way deterministic sensitivity analysis of the provider unit cost.
- **S1 Table**. Stage-specific cumulative costs of incomplete IVF cycles.
- **S2 Table**. Provider unit cost per completed IVF cycle and stage-specific cost distribution at MNH.
- **S3 Table**. One-way deterministic sensitivity analysis of major cost drivers for a completed IVF cycle at MNH.

